# Prolonged deployed hospital care in the management of military eye injuries

**DOI:** 10.1101/2020.06.27.20141648

**Authors:** Amrit Singh Dhillon, Mohammad Salman Zia Ahmad, John Breeze, Richard James Blanch

## Abstract

**Background/Objectives:** Prolonged hospital care is described as deployed medical care, applied beyond doctrinal planning timelines and military medical planning envisages that in future conflicts, patients will have to be managed for up to five days without evacuation to their home country. We aimed to investigate the effect of prolonged hospital care on visual outcomes in the management of open and closed globe injures.

**Methods:** We conducted a retrospective cohort study in the setting of British military operations in Afghanistan. We included consecutive UK military patients with ocular trauma evacuated from Afghanistan between December 2005 and April 2013. We assessed outcome using best corrected visual acuity (VA) 6-12 months after injury.

**Results:** All patients were male, with a mean age of 25. Outcomes adjusted for ocular trauma score (OTS) at presentation were similar to previous reports of military ocular trauma. The mean time to arrival at a centre with an ophthalmologist was 1.74 days. Both patients with penetrating open globe injuries and patients with hyphaema and an OTS of 3 or less displayed an association between worsening 6-12 month VA and time between injury and repair or assessment by an ophthalmologist.

**Conclusion:** Time to specialist ophthalmic care contributes to outcome after military open and closed globe injuries, supporting deployment of ophthalmologists on military operations.

## INTRODUCTION

Military ocular trauma is common, being found in 10% of all injuries from explosive devices on the modern battlefield. This high frequency is compounded by a high probability of long-term profound visual loss, reflecting the lethality of weaponry used in modern conflicts.^1, 2^ Unlike civilian ocular trauma which is predominantly unilateral, up to 37% of military eye injuries are bilateral.^1, 3-6^ Military injuries are more severe than civilian injuries, with up to half of all injuries affecting the retina, compared to less than 1% of civilian injuries. In addition, 5-7% of civilian open globe injuries require eye removal compared to 31% of military open globe injuries.^1, 3, 4, 7, 8^

Medical support to recent conflicts in Iraq and Afghanistan has relied on rapid aeromedical casualty evacuation, both to a medical treatment facility (MTF) with specialist surgeons (such as ophthalmologists) within the theatre of operations (Role 3), or out of country such as to Germany or the UK (Role 4). In the case of US support to Operations in Iraq (IRAQI FREEDOM and NEW DAWN) and Afghanistan (ENDURING FREEDOM and FREEDOM’S SENTINEL), this necessitated deploying Ophthalmologists to Iraq, Afghanistan and Germany for the entire conflicts,^9^ thus enabling assessment of eye injuries and repair of open globes within hours of injury – a level of care comparable to that found in civilian medical facilities.^1, 10, 11^ In the case of medical support to the United Kingdom (UK) Operation HERRICK in Afghanistan, Role 2 enhanced facilities in country relied on non-specialist initial assessment and stabilization, then rapid evacuation to a Role 4 facility in the UK,^7, 12^ which meant that between 2004 and 2008, the mean time taken for UK service personnel to be evacuated to specialist ophthalmic input (UK Role 4) was 2.6 days.^7^

NATO joint medical doctrine is to undertake damage control surgery within two hours of wounding.^13^ Although time to specialist treatment of ocular injuries has not been defined, multi-disciplinary consensus states that this should be within 24 hours.^14^ In a theatre of conflict, this level of medical support requires both the deployment of ophthalmologists and overall air-superiority.^11^ Future conflicts are likely to have delays in evacuation,^14,15^ and are likely to require local, Role 3, management for up to five days post injury.

The British military use the NATO doctrinal term “prolonged hospital care” (PHC) to describe “in theatre’ surgery” that is required when evacuation timelines are protracted.^13, 16^ The US military instead use the term prolonged field care (PFC) in Joint US doctrine as “field medical care, applied beyond doctrinal planning timelines”, and culminates in evacuation to higher level medical treatment facilities (MTF).^17^ For trauma, PHC and PFC may be thought of as an extension or follow-on treatment to Tactical Combat Casualty Care (TCCC), when evacuation is delayed and providers are forced to address the patient’s needs beyond the initial resuscitation and preparation for transport.^16^

The 2.6 day evacuation time previously reported therefore represents care beyond current multidisciplinary consensus and standard western practice and most servicemen in this series therefore form an example of the effect of prolonged field care of ophthalmic injuries.^3, 4, 18^ We aimed to determine any effects on visual outcomes of this prolonged time to specialist ophthalmic assessment and treatment of eye injuries in UK service personnel injured in Afghanistan.

## METHODS

The study was approved by the Clinical Governance departments of University Hospitals Birmingham NHS Foundation Trust (UHBFT) and Sandwell and West Birmingham NHS Trust (SWBH) and Research and Innovation at the Royal Centre for Defence Medicine (RCDM).

We performed a retrospective comparative cohort study. We included all consecutive cases of military deployment-related eye trauma presenting to the United Kingdom Role 4 MTF between 23 August 2007 and 19 April 2013. We collected clinical data from UHBFT (the major military trauma unit in the UK, including RCDM) and the Birmingham and Midland Eye Centre (a major tertiary referral unit, part of SWBH), to which some patients were transferred for treatment. As UK military policy during this period was for all aeromedical evacuations for deployed ophthalmic trauma to be transferred to RCDM, this included all cases of military ophthalmic trauma during that period.

Cases were identified from: the UK Joint Theatre Trauma Registry; searching diagnosis codes at UHBFT for diagnosis codes of ocular trauma in military patients; emergency operating room registers at SWBH; a prospectively collected register of all cases with head and neck injury as previously reported.^19^ Data were collected between July 2017 and July 2019. We recorded: patient demographics, mechanism and classification (Birmingham Eye Trauma Terminology System) of injury, presenting best-corrected visual acuity (VA), the presence of an afferent pupillary defect or traumatic infective endophthalmitis, surgical procedures performed (including primary repair and any secondary surgical interventions) and visual outcomes as best corrected visual acuity (VA). To mitigate inconsistencies associated with variable follow-up intervals, we recorded best corrected VA between 6 and 12 months post injury using the closest measurement to 6 months in instances where more than result was recorded. For the purposes of analysis, all VA measurements were converted to logarithm of the minimum angle of resolution (logMAR) equivalents, including those with less than Snellen vision, as previously described.^20^ Ocular trauma score was calculated retrospectively when there was sufficient documentation of injury type including: initial VA, presence or absence of a relative afferent pupillary defect, retinal detachment and endophthalmitis.^21^ Eye injuries were classified according to the Birmingham Eye Trauma Terminology System (BETTS).^22^ Secondary procedures were considered as any surgical procedure, directly related to the trauma, performed after the time of primary repair.

### Statistical analysis

Statistical analyses were performed in SPSS 21 (IBM, Armonk, NY, USA). Unless otherwise stated, means are given with standard error of the mean in brackets afterwards. Means were compared using student’s t-test. To account for the effects of injury severity on outcome, data were analysed using generalized linear models including all collected factors and covariates with categories with fewer than 7 cases combined and terms with a significance greater than p=0.05 or preventing model fit sequentially removed from the model. To account for the effect of missing data, sensitivity analysis used a chained equations method to impute five datasets for analysis of the effect of missing values.

## RESULTS

All patients were male, with a mean age of 25. Injuries are summarized in Figure 1. The mean time to arrival at a centre with an ophthalmologist was 1.74 days (±0.15; median 1.32; range 0.13 – 7.21). Outcomes, summarised by presenting ocular trauma score where this could be calculated are presented in Table 1. There was no variation in time to arrival by open or closed injury type.

**Table 1.** Outcome for all injuries with complete data broken down by initial and final VA and presenting ocular trauma score.

| Initial VA<br>/ OTS \ Final VA | >6/12 | 6/15-6/60 | 5/60-6/1200 | HM/LP | NPL |
| --- | --- | --- | --- | --- | --- |
| >6/12 | 37 | 1 | 2 | 2 | 0 |
| 6/15-6/60 | 11 | 1 | 0 | 0 | 0 |
| 5/60-6/1200 | 8 | 2 | 0 | 1 | 1 |
| HM/LP | 5 | 4 | 4 | 4 | 5 |
| NPL | 0 | 0 | 0 | 0 | 3 |
| OTS 5 | 9 | 0 | 0 | 0 | 0 |
| OTS 4 | 3 | 1 | 0 | 0 | 0 |
| OTS 3 | 6 | 4 | 2 | 2 | 1 |
| OTS 2 | 0 | 1 | 0 | 1 | 1 |
| OTS 1 | 0 | 0 | 0 | 0 | 3 |

**Figure 1.**
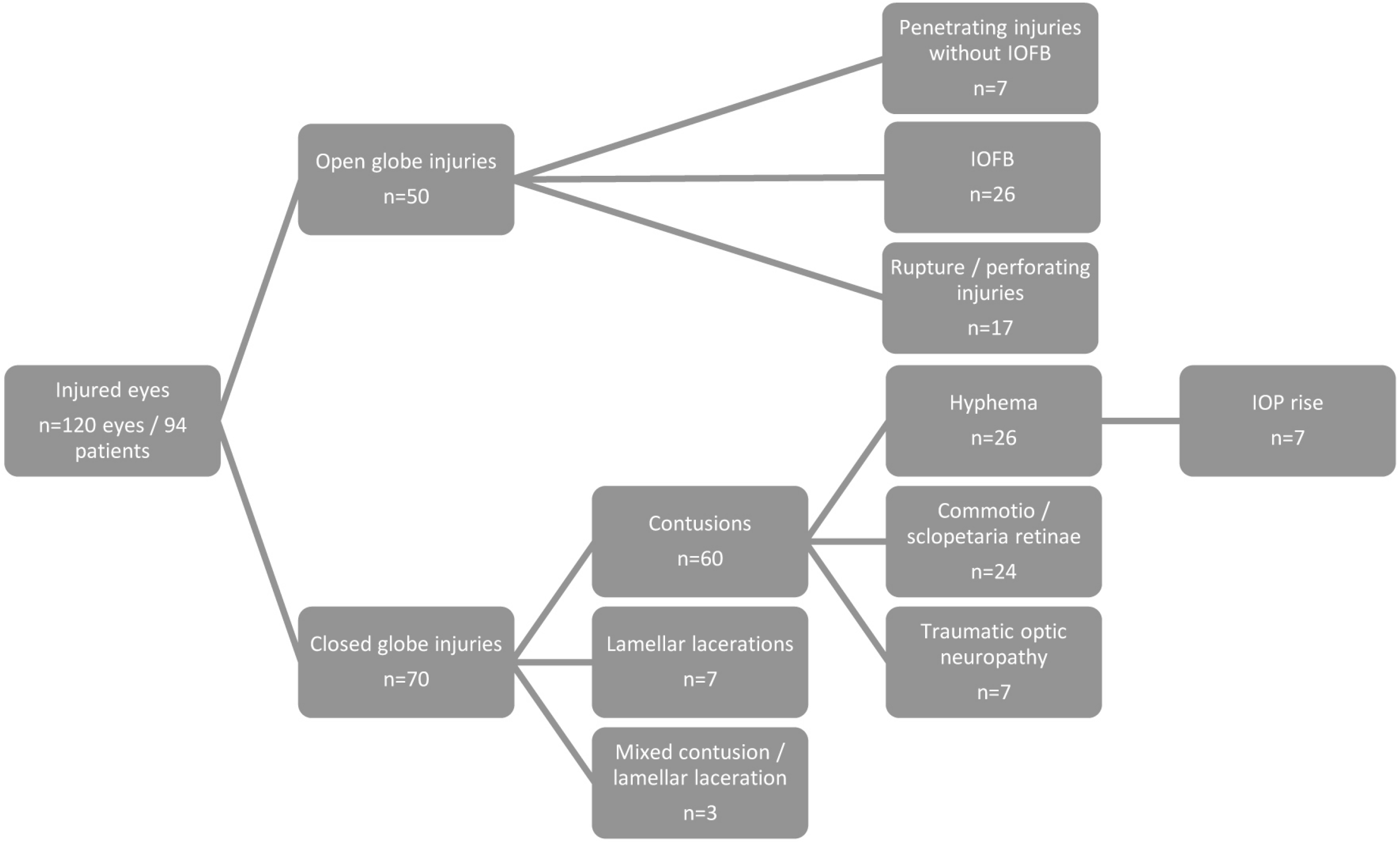
Summary of injury classification.

### OPEN GLOBE INJURIES

Because of the low number of perforating injuries and the frequent difficulty of distinguishing between these two types of military injury, rupture was classified with perforating injury for the purposes of analysis. One injury was a gunshot wound (perforating) and 49 were blast injuries. The mean time to arrival at a centre with an ophthalmologist was 1.34 days (±0.1).

#### Predictors of outcome after primary repair

Visual acuity 6-12 months after injury was available in 49/50 cases. BETTS, but not OTS, was independently associated with 6-12 month VA (Table 2). The mean time to primary open globe repair was 1.82 days (median 1.46, range 0.21-6.0). Increasing time to repair was associated with worse visual outcomes in penetrating (but not rupture or IOFB) injuries (Table 2). Sensitivity analysis to test the effect of missing data yielded results equivalent to the main analysis.

**Table 2.** Visual acuity (VA) outcomes 6-12 months after injury in open and closed globe injuries and the effect of time from injury to primary repair or specialist ophthalmic care for each injury type. P values relate to the effect of time to specialist ophthalmic care on 6-12 month VA, with effects having a p value <0.05 shown in bold.

| Injury classification | Mean 6-12 month logMAR VA | Increase in 6-12 month logMAR VA per day to specialist ophthalmic care | p value |
| --- | --- | --- | --- |
| Penetrating injury | 2.14 ($\pm 0.77$) | **3.18 ($\pm 1.18$)** | **0.007** |
| IOFB | 1.49 ($\pm 0.33$) | -0.37 ($\pm 0.34$) | 0.287 |
| Rupture/perforating injury | 4 ($\pm 0$) | 0 ($\pm 0.25$) | 1 |
| Hyphaema with OTS$\leq 3$ | 1.16 ($\pm 0.25$) | **1.27 ($\pm 0.42$)** | **<0.001** |
| Hyphaema with OTS$\geq 4$ | 0.03 ($\pm 0.05$) | 0 ($\pm 0.17$) | 0.893 |

#### Endophthalmitis

There were three cases of endophthalmitis, two penetrating injuries and one with IOFB, which had times to repair of 1.39, 1.68 and 2.0 days and were all secondarily eviscerated within one month of injury. When these cases were excluded from the analysis of the effect of time to repair, the effect of increasing time to repair on penetrating injuries was relatively unchanged at logMAR 3.12 (±1.12) per 24 hours.

#### Other procedures

Twenty one patients had vitrectomy, a median of 4 days (range 0-26) after primary repair, of whom one went on to have secondary evisceration. Seven patients had primary evisceration, three in theatre (two by ophthalmologists in US facilities in theatre) and four on return to the UK. Fourteen patients had secondary eviscerations and two had secondary enucleations, a median of 21 days after primary repair (range 4-321).

### CLOSED GLOBE INJURIES

Visual outcomes for the different injury types and factors affecting outcome are summarized in Table 2. The mean time to arrival at a centre with an ophthalmologist was 1.82 days (±0.28). Initial size of hyphaema was not documented except in one case when it was total and the patient subsequently experienced an IOP rise.

Patients with hyphaema had worse 6-12 month VA (0.63±0.20) than patients who did not (0.12±0.06; p=0.047), despite accounting for injury severity using OTS. In patients with hyphaema and severe injury, indicated by OTS of 3 or less, increasing time to ophthalmic care was associated with a decrease in 6-12 month visual acuity (Table 2).

Because initial OTS was missing in 19/70 cases and 6-12 month VA in 7/70 cases, we performed a sensitivity analysis with multiple imputation of the missing values, in which the effect of time to ophthalmic care did not remain significant in the model.

### BILATERAL INJURY

Eight patients had bilateral open globe injury. Including closed globe injuries, 28 patients had bilateral injury and three patients had worse than Snellen acuity in both eyes at final follow up; one patient had bilateral eviscerations (one primary, one secondary), one patient had a final vision of perception of light in his better eye and one could count fingers in his better eye.

## DISCUSSION

PHC includes care provided outside of doctrinal planning timelines. We report the care of deployed military eye injuries, most of which received initial ophthalmic care more than 1 day after injury, ranging up to 7 days. While visual outcomes were comparable with other military series,^1^ both closed and open globe injuries suffered decrements in final visual acuity with increasing time to specialist assessment and the probability of endophthalmitis was increased compared to comparable military injuries with more rapid access to care.^1, 10^

The mean time to specialist ophthalmic assessment was 1.5 days, which is less than previously reported and represents an incredible achievement of the aeromedical evacuation process.^7^ The standard of care in Western medical practice and consensus for military injury management is access to specialist assessment to allow primary repair within 24 hours of injury and there is some evidence that this should be shortened to 12 hours.^18^ Delayed repair beyond 12-24 hours worsens final visual acuity and increases the risk of endophthalmitis and post-operative wound leak,^3, 4, 18, 23^ consistent with our finding of worsening visual outcome associated with time to repair for penetrating injuries. The lack of any association in rupture and perforating injuries relates to the universally poor outcome for this injury type (all injuries no perception of light or eviscerated at 6-12 months). The lack of association for IOFB injuries could reflect development of cataract after secondary vitreoretinal surgery. IOFB injuries being lower energy (with better visual outcome) is an unlikely explanation, as that is the case in civilian injuries, in which an association with time to repair is present.^3^

OTS was not associated with visual outcome when BETTS was also included in the analysis, which most probably reflects a lack of independence between these two terms, with rupture injuries associated with a low OTS for instance.

When compared with civilian injuries in Birmingham in a similar timeframe,^3^ military injuries had worse outcomes even accounting for the effect of OTS, BETTS injury type and time to surgery with military patients’ 6-12 month logMAR VA 0.96 (±0.36) worse than civilians’. Compared to the analysis of 253 globe injuries by Weichel et al.,^1^ broken down by presenting visual acuity, there was no evidence of a difference in outcome compared to our series (Chi-squared, p=0.496), but endophthalmitis was more common (Chi-squared, p<0.001).

Three cases of endophthalmitis from 50 open globe injuries is similar to civilian series,^3, 24, 25^ in which times to primary repair were variable (but lower than this series) and antibiotics were not always given, but higher than comparable military case series, in which systemic antibiotic prophylaxis was given and primary repair performed within hours of injury,^1, 10^ suggesting that earlier access to primary repair reduces the risk of endophthalmitis even when systemic antibiotic prophylaxis is given.

For closed injuries, delayed access to care in hyphaema (>24 hours) is associated with an increased re-bleeding rate.^26^ We observed worse visual outcomes for hyphaema patients compared to other patients with closed globe injury, despite accounting for injury severity using OTS, which could suggest that the presence of hyphaema is associated with a more severe injury than indicated by reductions in presenting visual acuity or that the complications of hyphaema worsen visual outcome. The effect of time to ophthalmic care in patients with hyphaema and OTS ≤3 suggests that patients with hyphaema and more severe injuries were more likely to suffer complications reducing visual outcome, such as rebleed and elevated IOP, and the likelihood of this increased with increased time to ophthalmic care, which would be consistent with previous evidence.^26^

We recognize multiple limitations to this study, which, like other analyses of military ocular trauma, was retrospective in nature. We have attempted to address variation in injury severity by calculation of OTS, but are limited by the proportion of patients in whom visual acuity could not be assessed because of unconsciousness. We have further accounted for potential variation in injury severity in different groups by sensitivity analysis to impute missing OTS values, which found results consistent with the primary analysis in the case of open globe injuries, supporting a reliable association between time to primary open globe repair and visual outcome.

The potential for PHC in future conflicts remains a challenge for military medical planners, in which there are three options for military ophthalmic injury management: (1), forward deployment of ophthalmology; (2), delayed access to ophthalmology; (3), ophthalmic care provided by non-specialists. This cohort includes patients managed under a combination of all three approaches, as some patients were assessed and managed within hours of injury by US Ophthalmologists in country, some had limited assessment and treatment by non-specialists and most waited for specialist ophthalmic assessment in the UK. These data suggest that time to specialist ophthalmic care contributes to outcome and should support forward deployment of ophthalmology.

## Data Availability

All necessary data is contained in the manuscript.

## REFERENCES

1. Weichel ED, Colyer MH, Ludlow SE, Bower KS, Eiseman AS. Combat ocular trauma visual outcomes during operations iraqi and enduring freedom. Ophthalmology 2008; 115(12): 2235–2245.

2. Wong TY, Klein BE, Klein R. The prevalence and 5-year incidence of ocular trauma. The Beaver Dam Eye Study. Ophthalmology 2000; 107(12): 2196–2202.

3. Blanch RJ, Bishop J, Javidi H, Murray PI. Effect of time to primary repair on final visual outcome after open globe injury. Br J Ophthalmol 2019; 103(10): 4.

4. Kong GY, Henderson RH, Sandhu SS, Essex RW, Allen PJ, Campbell WG. Wound-related complications and clinical outcomes following open globe injury repair. Clin Exp Ophthalmol 2015; 43(6): 508–513.

5. Negrel AD, Thylefors B. The global impact of eye injuries. Ophthalmic Epidemiol 1998; 5(3): 143–169.

6. Blanch RJ, Good PA, Shah P, Bishop JR, Logan A, Scott RA. Visual outcomes after blunt ocular trauma. Ophthalmology 2013; 120(8): 1588–1591.

7. Blanch RJ, Bindra MS, Jacks AS, Scott RA. Ophthalmic injuries in British Armed Forces in Iraq and Afghanistan. Eye 2011; 25(2): 218–223.

8. Jones NP, Hayward JM, Khaw PT, Claoue CM, Elkington AR. Function of an ophthalmic “accident and emergency” department: results of a six month survey. Br Med J (Clin Res Ed) 1986; 292(6514): 188–190.

9. Blanch RJ, Kerber MT, Gensheimer WG. Deployed ophthalmic workload in support of US and NATO operations in Afghanistan. BMJ Mil Health 2020; epub 5 Mar 2020.

10. Colyer MH, Weber ED, Weichel ED, Dick JS, Bower KS, Ward TP et al. Delayed intraocular foreign body removal without endophthalmitis during Operations Iraqi Freedom and Enduring Freedom. Ophthalmology 2007; 114(8): 1439–1447.

11. Breeze J, Blanch R, Mazzoli RA, Bowley DM, DuBose JJ, Rickard R et al. Comparing the management of eye injuries by deployed US and UK military surgeons during the Iraq and Afghanistan conflicts. Ophthalmology 2019; S0161-6420: 32148–32147.

12. Maitland L, Lawton G, Baden J, Cubison T, Rickard R, Kay A et al. The Role of Military Plastic Surgeons in the Management of Modern Combat Trauma: An Analysis of 645 Cases. Plast Reconstr Surg 2016; 137(4): 717e–724e.

13. Allied Joint Doctrine for Medical Support. B ed: Ministry of Defence; 2015:1–280.

14. Breeze J, Blanch R, Baden J, Monaghan AM, Evriviades D, Harrisson SE et al. Skill sets required for the management of military head, face and neck trauma: a multidisciplinary consensus statement. J Roy Army Med Corps 2018; 164(2): 133–138.

15. Soliz BA. Saving Lives in Prolonged Care Scenarios at Role 1 Requires Changes in Leadership, Force Structure, and Training: US Army War College Carlise United States; 2018.

16. DeSoucy E, Shackelford S, DuBose JJ, Zweben S, Rush SC, Kotwal RS et al. Review of 54 Cases of Prolonged Field Care. J Spec Oper Med 2017; 17(1): 121–129.

17. Keenan S. Deconstructing the Definition of Prolonged Field Care. J Spec Oper Med 2015; 15(4): 125.

18. Essex RW, Yi Q, Charles PGP, Allen PJ. Post-traumatic endophthalmitis. Ophthalmology 2004; 111(11): 2015–2022.

19. Breeze J, McVeigh K, Lee JJ, Monaghan AM, Gibbons AJ. Management of maxillofacial wounds sustained by British service personnel in Afghanistan. Int J Oral Maxillofac Surg 2011; 40(5): 483–486.

20. Schulze-Bonsel K, Feltgen N, Burau H, Hansen L, Bach M. Visual acuities “hand motion” and “counting fingers” can be quantified with the freiburg visual acuity test. Invest Ophthalmol Vis Sci 2006; 47(3): 1236–1240.

21. Kuhn F, Maisiak R, Mann L, Mester V, Morris R, Witherspoon CD. The Ocular Trauma Score (OTS). Ophthalmol Clin North Am 2002; 15(2): 163–165, vi.

22. Kuhn F, Morris R, Witherspoon CD. Birmingham Eye Trauma Terminology (BETT): terminology and classification of mechanical eye injuries. Ophthalmol Clin North Am 2002; 15(2): 139–143, v.

23. Cruvinel Isaac DL, Ghanem VC, Nascimento MA, Torigoe M, Kara-Jose N. Prognostic factors in open globe injuries. Ophthalmologica 2003; 217(6): 431–435.

24. Thompson JT, Parver LM, Enger CL, Mieler WF, Liggett PE. Infectious endophthalmitis after penetrating injuries with retained intraocular foreign bodies. National Eye Trauma System. Ophthalmology 1993; 100(10): 1468–1474.

25. Thompson WS, Rubsamen PE, Flynn HW, Schiffman J, Cousins SW. Endophthalmitis after Penetrating Trauma - Risk-Factors and Visual-Acuity Outcomes. Ophthalmology 1995; 102(11): 1696–1701.

26. Fong LP. Secondary hemorrhage in traumatic hyphema. Predictive factors for selective prophylaxis. Ophthalmology 1994; 101(9): 1583–1588.

